# Identification of super-transmitters of SARS-CoV-2

**DOI:** 10.1101/2020.04.19.20071399

**Authors:** Xuemei Yang, Ning Dong, Edward Wai-Chi Chan, Sheng Chen

**Author notes:** Corresponding author: Sheng Chen, City University of Hong Kong, Kowloon, Hong Kong. contribute equally to the work (Xuemei Yang, Ning Dong).

## Abstract

A newly emerged coronavirus, SARS-CoV-2, caused severe outbreaks of pneumonia in China in December 2019 and has since spread to various countries around the world. To probe the origin and transmission dynamics of this virus, we performed phylodynamic analysis of 247 high quality genomic sequences of viruses available in the GISAID platform as of March 05, 2020. A substantial number of earliest sequences reported in Wuhan in December 2019, including those of viruses recovered from the Huanan Seafood Market (HNSM), the site of the initial outbreak, were genetically diverse, suggesting that viruses of multiple sources were involved in the original outbreak. The viruses were subsequently disseminated to different parts of China and other countries, with diverse mutational profiles being recorded in strains recovered subsequently. Interestingly, four genetic clusters defined as Super-transmitters (STs) were found to become dominant and were responsible for the major outbreaks in various countries. Among the four clusters, ST1 is widely disseminated in Asia and the US and mainly responsible for outbreaks in the states of Washington and California in the US as well as those in South Korea at the end of February and early March, whereas ST4 contributed to the pandemic in Europe. Each ST cluster carried a signature mutation profile which allowed us to trace the origin and transmission patterns of specific viruses in different parts of the world. Using the signature mutations as markers of STs, we further analysed 1539 genome sequences reported after February 29, 2020. We found that around 90% of these genomes belonged to STs with ST4 being the dominant one and their contribution to pandemic in different continents were also depicted. The identification of these super-transmitters provides insight into the control of further transmission of SARS-CoV-2.

## Introduction

A number of newly emerged coronaviruses such as the highly pathogenic severe acute respiratory syndrome coronavirus (SARS-CoV) and Middle East respiratory syndrome coronavirus (MERS-CoV) have caused serious respiratory and intestinal infections in human within the past two decades [1]. In December 2019, another new coronavirus, SARS-CoV-2, has emerged and caused outbreaks of lower respiratory tract infections, often with poor clinical outcome, in Wuhan, China. The virus, which has since spread to other cities in China and various countries worldwide [2], exhibited a high potential to undergo human-to-human transmission [3]. As of March 26, 2020, a total of 692 thousand infections were documented worldwide, among which 68 thousand occurred in China (https://www.gisaid.org/epiflu-applications/global-cases-betacov/). As a result, WHO declared the risk of SARS-CoV-2 as “Very High” in China (https://www.who.int/docs/default-source/coronaviruse/situation-reports/20200202-sitrep-13-ncov-v3.pdf).

The genomic characteristics of SARS-CoV-2 have been elucidated using phylogenetic, structural and mutational analyses by scientists across the globe [4]. High-throughput sequencing revealed that SARS-CoV-2 was a novel betacoronavirus which resembled SARS-CoV at around 79.5% sequence identity [5, 6]. A recent study indicated that SARS-CoV-2 was 96% identical to a bat coronavirus RaTG13 (accession: MN996532) at the genomic level, suggesting that bat might be a natural host of SARS-CoV-2 [7]. GISAID is a platform for sharing genetic data of influenza. Currently, a rapidly increasing number of SARS-CoV-2 genomic sequences are being deposited into this database from laboratories around the world [8]. On the other hand, some recent studies also suspected that Malayan pangolins (*Manis javanica*) could be the intermediate host of this new coronavirus, since the amino acid sequence of the S protein of coronaviruses derived from Malayan pangolins illegally imported to Guangdong Province of China, as well as coronaviruses harboured by pangolins in Guangxi province of China, exhibited very high homology with the S protein sequence of SARS-CoV-2, even though the overall homology between SARS-CoV-2 and RaTG13 is still the highest[9]. However, due to the inability to detect or isolate SARS-CoV-2 from pangolins in the Wuhan Huanan seafood wholesale market, the site in which the first batch infected patients had commonly visited, the theory of pangolins being the culprit of the Wuhan pneumonia outbreak is not substantiated. The intermediate host for SARS-CoV-2 therefore remains a mystery. In fact, it remains unclear if the Huanan Seafood Wholesale Market is the origin of this outbreak as some of the earliest cases were confirmed to have no linkage with this market. It is urgent to identify the source(s) of viruse(s) that caused this outbreak to design more effective control measures to stop the continuous worldwide transmission of these highly contagious viruses. With more sequences released, it is very important to provide more insights of this virus through indepth sequence analysis. One recent study has analysed over 100 available genome sequences and revealed that sequences belonging to different genetic clusters have evolved [10]. In this study, we retrieved and analysed the publicly shared genome sequences as of March 25, 2020 to investigate the genetic diversity and phylodynamics of these SARS-CoV-2 viruses. We identified notable four clusters of genomes with high transmission and mutation rate and have become the most dominant viruses in the later stage of transmission. Results in this study should provide insight into the control of further transmission of SARS-CoV-2.

## Materials and Methods

### Sequence analysis, alignment and mutation identification

A total of 343 full-length SARS-CoV-2 genomes available in the GISAID platform (https://platform.gisaid.org/) as of March 5, 2020 were downloaded [8]. A total of 247 sequences with high sequence quality as noted in the GISAID database were included for further analysis after removing sequences containing little temporal signal and thus are not unsuitable for inference using phylogenetic molecular clock models. Information regarding the date and country of isolation were also retrieved from the GISAID platform. The annotated reference genome sequence of the SARS-CoV-2 isolate Wuhan-Hu-1 (accession: NC_045512.2) was downloaded from the NCBI GenBank database. All genomes were annotated by GATU Genome Annotator [11] using the SARS-CoV2 isolate Wuhan-Hu-1 (NC_045512.2) as reference [12]. Nucleotide and amino acid mutations of all genome and separate proteins were analyzed by blast (https://blast.ncbi.nlm.nih.gov/) using the sequence of strain Wuhan-Hu-1 as reference.

### Phylodynamic analysis

Global genomic surveillance of SARS-CoV-2 was implemented by means of an automated phylogenetic analysis pipeline using Nextstrain, which generates an interactive visualization integrating a phylogeny with sample metadata such as geographic location or host age [13]. The pipeline involved the sequence alignment module with MAFFT [14], phylogenetic analysis with IQ-TREE [15], maximum-likelihood phylodynamic analysis with Treetime [16], identification of nucleotide and amino acid mutations with Augur, and result visualization with Auspice [13]. The outputs were edited by Inkscape 0.91 [17].

### Phylogenetic analysis

Alignment of the complete genome sequences was conducted with MAFFT v7.310 [14]. Phylogenetic tree of all SARS-CoV-2 was built with RAxML version 8.2.4 [18]. The tree was edited by iTOL [19].

### Quick identification of the types of SARS-CoV-2 genomes in the database

All complete genomes available as March 28 on the GISAID database were downloaded. Single Nucleotide Polymorphisms (SNPs) calling were performed by Snippy (https://github.com/tseemann/snippy) using Wuhan-Hu-1 as reference. Super-transmitter clusters were classified according relative variants. A total of 1956 qualified genomes submitted after February 29, 2020 were included.

## Results

### Phylodynamics analysis of genome sequences of SARS-CoV-2 strains collected worldwide

To trace the evolutional process and identify the common ancestor of 247 strains of SARS-CoV-2 collected worldwide, root-to-tip regression scatter plots was conducted among all SARS-CoV-2 genomes, with R^2^ being found to be 0.23, suggesting that these 247 viral sequences shared a common recent ancestor **(Fig 1a**). The date of the most recent common ancestor (tMRCA) of all reported SARS-CoV-2 viruses was 2019-Nov-12, suggesting that this virus emerged recently **(Fig 1a**). A total of 379 nucleotide mutations were identified among these 247 sequences based on the sequence alignment, among which G^11083^T (n=5), T3G (n=3), G^29864^A (n=3), C^29870^A (n=3), A1T (n=2), A4T (n=2), T^4402^C (n=2), G^5062^T (n=2), T^18603^C (n=2) and G^22661^T (n=2) were the most homoplasic mutations (Fig 2, supplementary file 1). A total of 147 strains were found to contain single amino acid change, with the majority of such changes being located within ORF1ab (n=104). The L^3606^F change was detected in two viral sequences, while other mutations occurred only once. Mutations that result in amino acid changes include single substitution in the S protein (n=19, D^614^G, L^752^F, F^32^I, H^655^Y, V^483^A, F^157^L, V^615^L, K^202^N, S^939^F, F^797^C, A^93^0V, R^408^I, V^367^F, Q^409^E, S^254^F, A435S, D^1146^E, S^247^R and P^1143^L), ORF3a (n=8, E^191^G, G^76^S, K^61^N, V^259^L, T^176^I, L^140^V, T^269^M and V^88^L), N protein (n=6, K^247^I, S^194^L, P^46^S, S^327^L, E^378^Q and D^343^V), ORF8 (n=4, T^11^I, L^84^S, S^97^N and S^67^F), ORF7a (n=3, P^34^S, Q62* and H^73^Q), ORF10 (n=2, P^10^S and I^13^M) and E protein (n=1, S^6^L) (**Fig 1b**). Identification of single amino acid substitutions in SARS-CoV-2 isolates consistently showed that these isolates shared a recent common ancestor but entered diverse evolution paths. The estimated substitution rate of SARS-CoV-2 was 8.90e-04 subs/site/year, which was similar to that of other RNA viruses including SARS-CoV, Ebola virus, Zika virus, and others, which was found to be at ∼ 1e-3 subs/site/year (http://virological.org/t/phylodynamic-analysis-93-genomes-15-feb-2020/356). Based on this mutation rate, a genome of 29kb of SARS-CoV-2 will end up with ∼26 mutations per genome per year, suggesting that within the two months’ study period, the number of mutations in each genome should not exceed five if all test isolates emerged as a result of natural evolution of a single SARS-CoV-2 strain.

**Figure 1.**
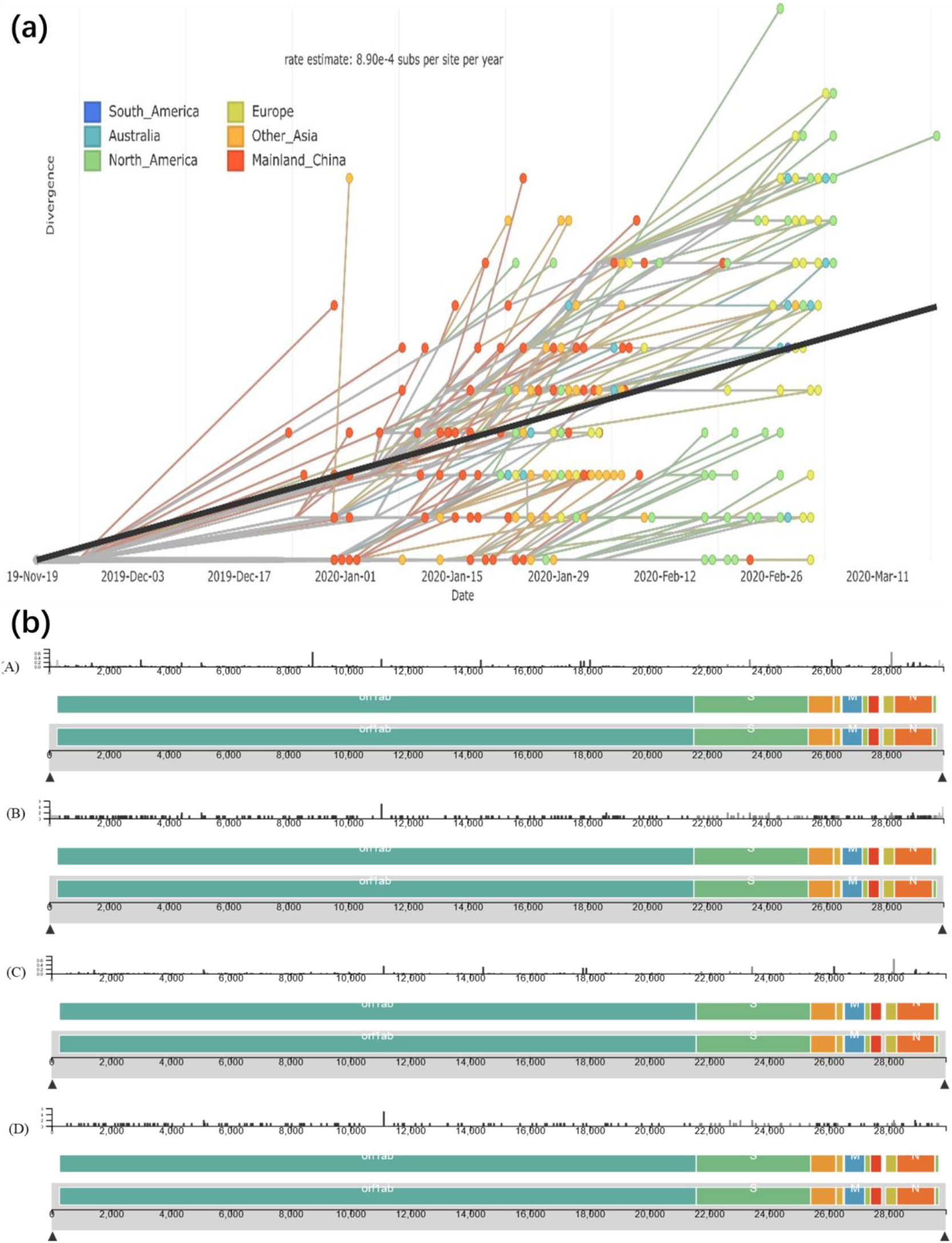
Root-to-tip regression scatter plots and distribution of mutations across the SAR-CoV-2 genomes. (a) Root-to-tip regression scatter plots of different strains of SARS-CoV-2. (b) Distribution of mutations across the SAR-CoV-2 genome. (A) and (B) illustrated the entropy and events of nucleotide mutations. (C) and (D) illustrated the entropy and events of amino acid mutations. Figure 1. Root-to-tip regression scatter plots of different strains of SARS-CoV-2. Dots in the plot indicate the SARS-CoV-2 isolates used in this study. The color of each dot represents the region of isolation of the corresponding isolate. Ancestral state reconstruction and branch length timing were performed with TreeTime [16].

**Figure 2.**
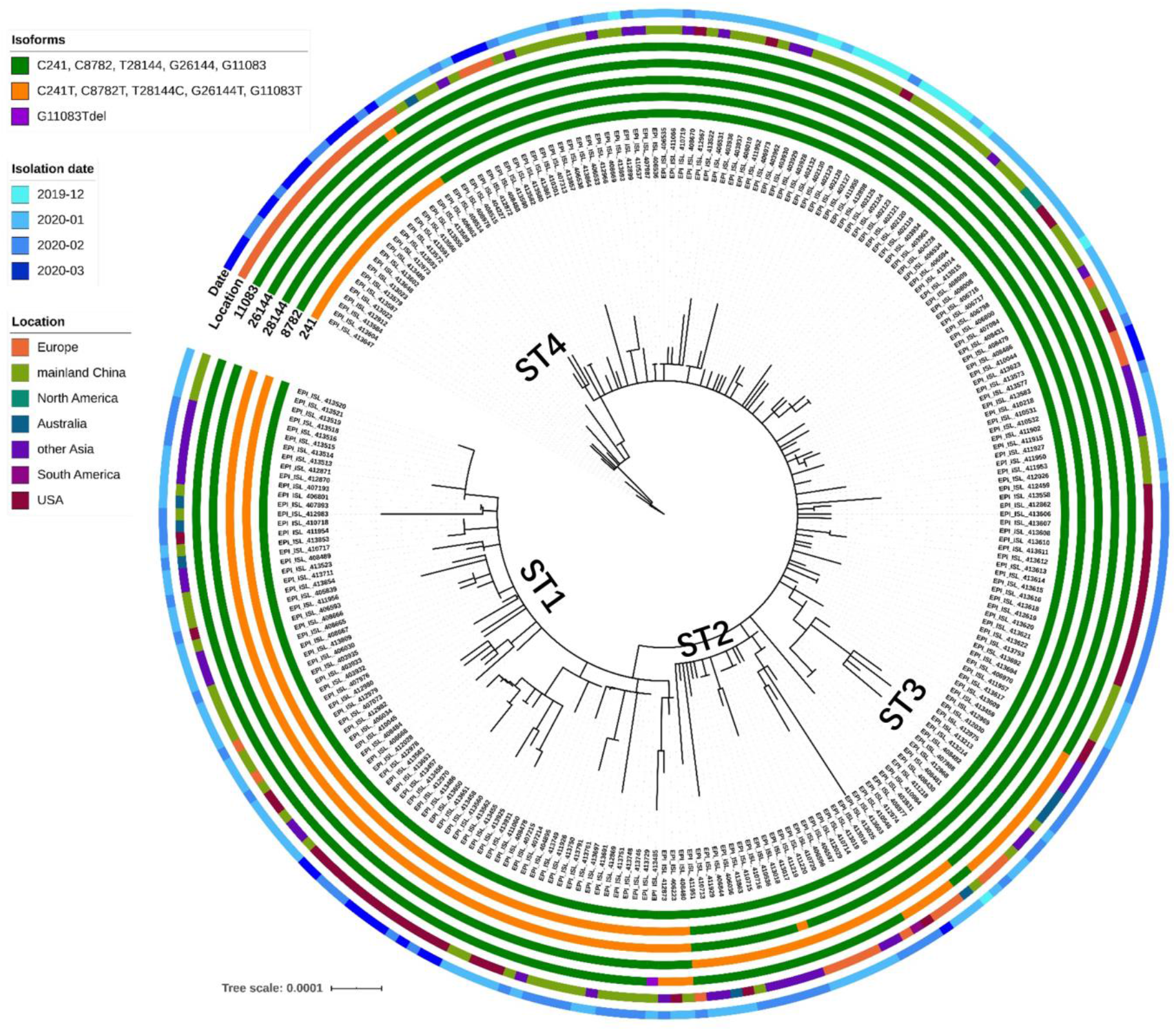
Phylogentic analysis of SARS-CoV-2 genomes. Four super-transmitter clusters (STs) were identified. Each ST was found to exhibit a signature mutation profile.

### Multiple origins of SARS-CoV-2 in Wuhan, China

To shed light on the evolution trend of SARS-CoV-2, we analysed the time-dependent changes in mutation profiles of the test strains in detail. A total of 16 viral genomes collected before Jan. 01, 2020, were included (**Table 1**). All of these 16 genomes were obtained from Wuhan, with half of them from Huanan Seafood Wholesale Market (HNSM) where the original outbreak occurred. Six genomes contained identical sequences, four of which belonged to isolates obtained from HNSM. Compared to these six viral genomes, others displayed various mutation profiles which comprised 1 to 6 mutations in the genomes. We therefore set these six genomes as reference genome for subsequent analyses. Two earliest viral genomes reported on Dec. 24 and 26 were found to harbour two and three mutations when compared to the reference viral genome, respectively. Four viral genomes from HNSM also contained two mutations with different profiles, suggesting that the original SARS-CoV-2 strain might have been circulating in HNSM for a certain period of time and underwent mutational changes in different intermediate hosts (**Table S1**). These observations suggested that HNSM was not the only origin of the COVID-19 outbreak, instead the market might only serve as a medium in which transmission of this virus to human first occurred. The original virus seemed to have transmitted to various provinces in China subsequently, including Guangdong, Zhejiang, Anhui, Jiangshu and Chongqing, and then to other countries including Japan, Taiwan, Thailand and USA in the following month (January 2020). The viral genome reported initially from USA were those of viruses recovered from the patients in Princess Diamond Cruise, confirming that the original virus was the one that caused the outbreak in this cruise; such view is consistent with the finding that identical genomes were reported in Japan, where the cruise ship was docked. A total of 26 out of the 247 genome sequences tested contain one mutation. Unless isolated from the same location, most of these genomes exhibit unique mutational profile. Five sequences from the Princess Diamond cruise ship were found to exhibit unique mutation profiles, thus further suggesting that random mutations occurred during viral evolution. It should be noted that these genome sequences were also reported in Wuhan, other parts of China and various other countries, confirming that the transmission of the original virus to different parts of the world was accompanied by active but random mutational changes during the process (**Table S1**).

**Table 1.**
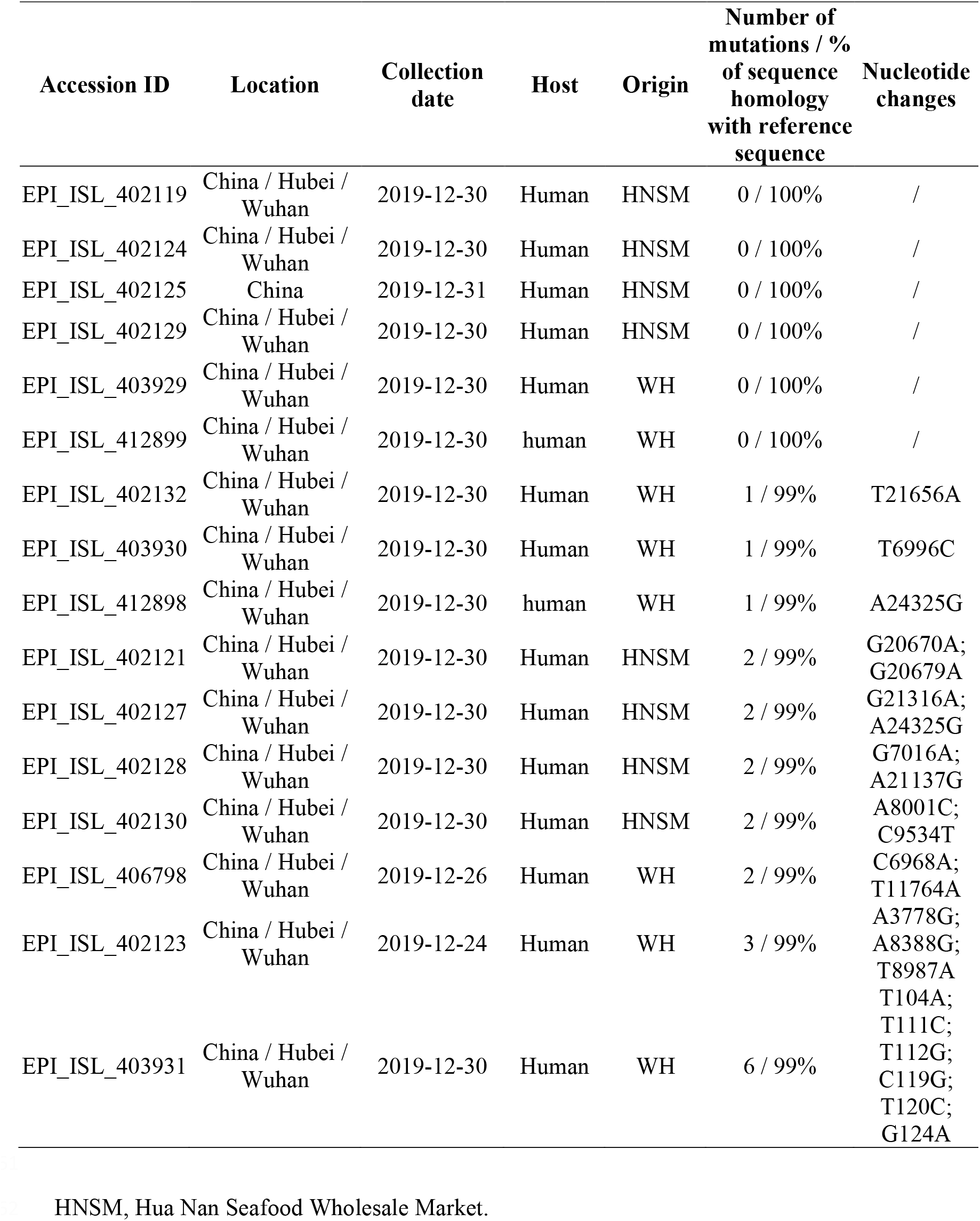
Mutational profile analysis of SARS-CoV-2 genome sequences obtained in December of 2019.

### Phylogenetic analysis of genome sequences of SARS-CoV-2

Phylogenetic analysis of the 247 SARS-CoV-2 genomes was also performed, with results showing that such viral genomes exhibited highly diverse genetic profiles and that random mutations occurred during the evolutional process within the first two months. Interestingly, four clusters of genome sequences were observed among the 247 genomes, with the rest exhibiting more diverse profiles. These results were consistent with the data of maximum-likelihood phylodynamic analysis shown in Figure 1. Comparison of the mutation profile of each cluster enabled us to discover that all viral genomes in the same cluster were derived from one parental viral genome sequence which bears a signature mutation profile, as such profile could be identified in all offsprings (**Fig 2**). The first cluster contained two mutations, C^8782^T and T^28144^C; the second cluster contained the mutation G^26144^T; the third cluster contained the mutation G^11083^T; the fourth cluster contained three mutations, C^241^T, C^3037^T and A^23403^G. Tracing the changes in mutation profiles of these viral genomes over time allowed us to visualize the transmission and evolution dynamics of SARS-CoV-2. Since viruses of all of these four clusters exhibited very high potential to undergo global transmission, we define viruses in these four clusters as super-transmitter cluster 1 (ST1), 2 (ST2), 3(ST3) and 4(ST4).

### Evolution and transmission of super-transmitter cluster 1 (ST1)

ST1 carried the signature mutation profile of C^8782^T and T^28144^C. The C^8782^T change is a silent mutation, whereas T^28144^C is associated with the amino acid substitution L^84^S in the Orf8ab protein. The ST1 viruses were transmitted very efficiently and a total of 85 out of 247 (34%) genome sequences belonging to this cluster as of March 03, 2010. The earliest sequence of in this cluster was reported in Wuhan, China on Jan 05, 2020 and seven were subsequently reported in January and February in different parts of China and Australia, suggesting that widespread transmission of this cluster of viruses occurred (**Table 2**). The viruses in ST1 were mainly transmitted among Asian countries area such as China, Japan, South Korea, Taiwan and Singapore, as well as North America, in particular the states of California and Washington in USA (**Table 2**). The viruses in ST1 were also found to be able to rapidly mutate along the transmission paths. Three genome sequences that were reported in Australia, Vietnam and USA on Feb. 28, 24 and Mar. 03, 2020 respectively, were found to harbour a total of 11 mutations. An additional nine mutations were acquired by the parental virus within 50 days (from Jan 05 to Feb. 24, 2020), with a mutation rate of 2.3e-3 subs/site/year (29kb genome size), which was much higher than the predicted mutation rate of SARS-CoV-2 (4.057 e-4 subs/site/year) and other coronaviruses such as SARS-CoV-1 and MERS virus. Among viral genomes in this cluster, 43 out of 85 genomes exhibited five or more mutations (**Table 2**).

**Table 2.**
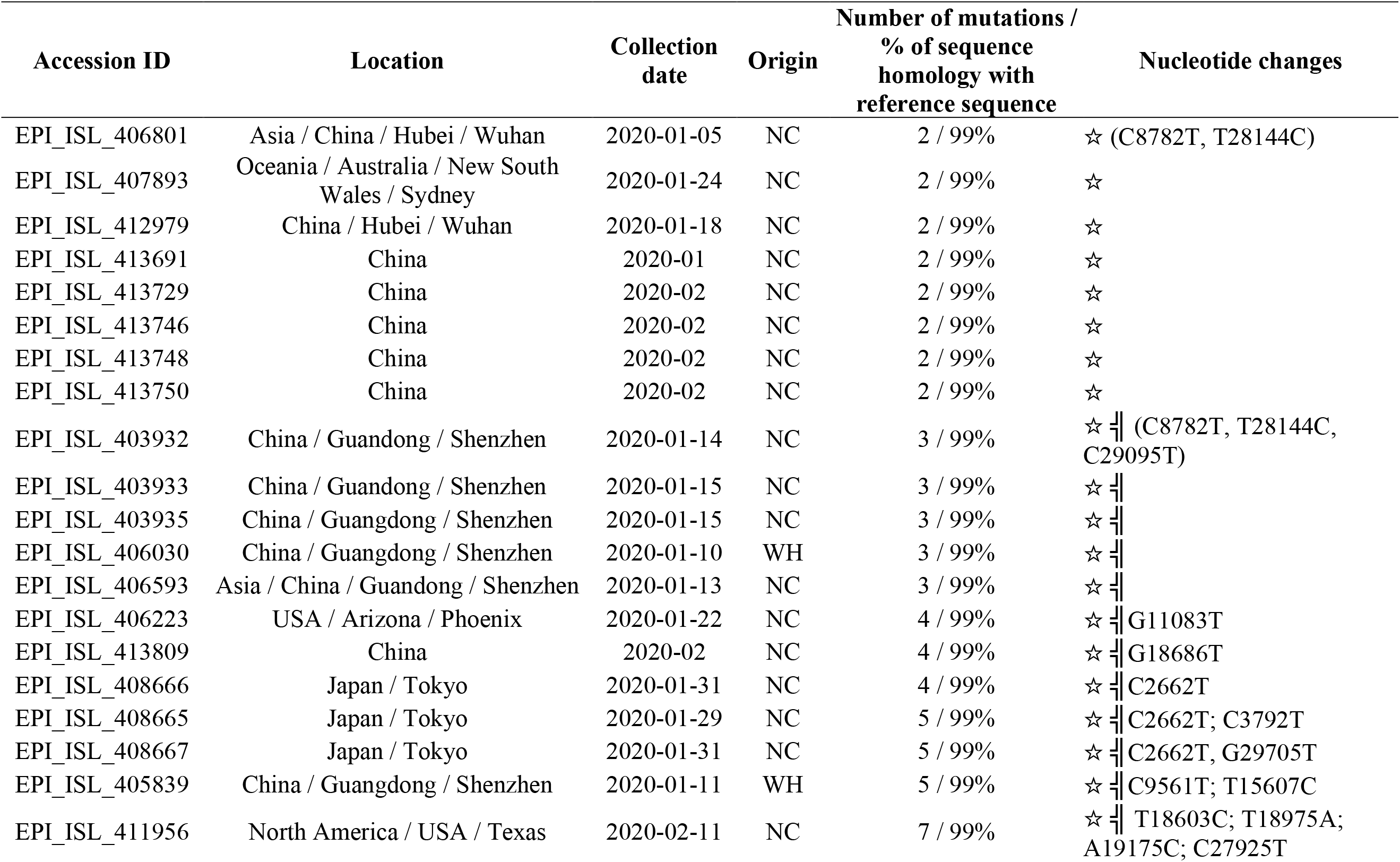
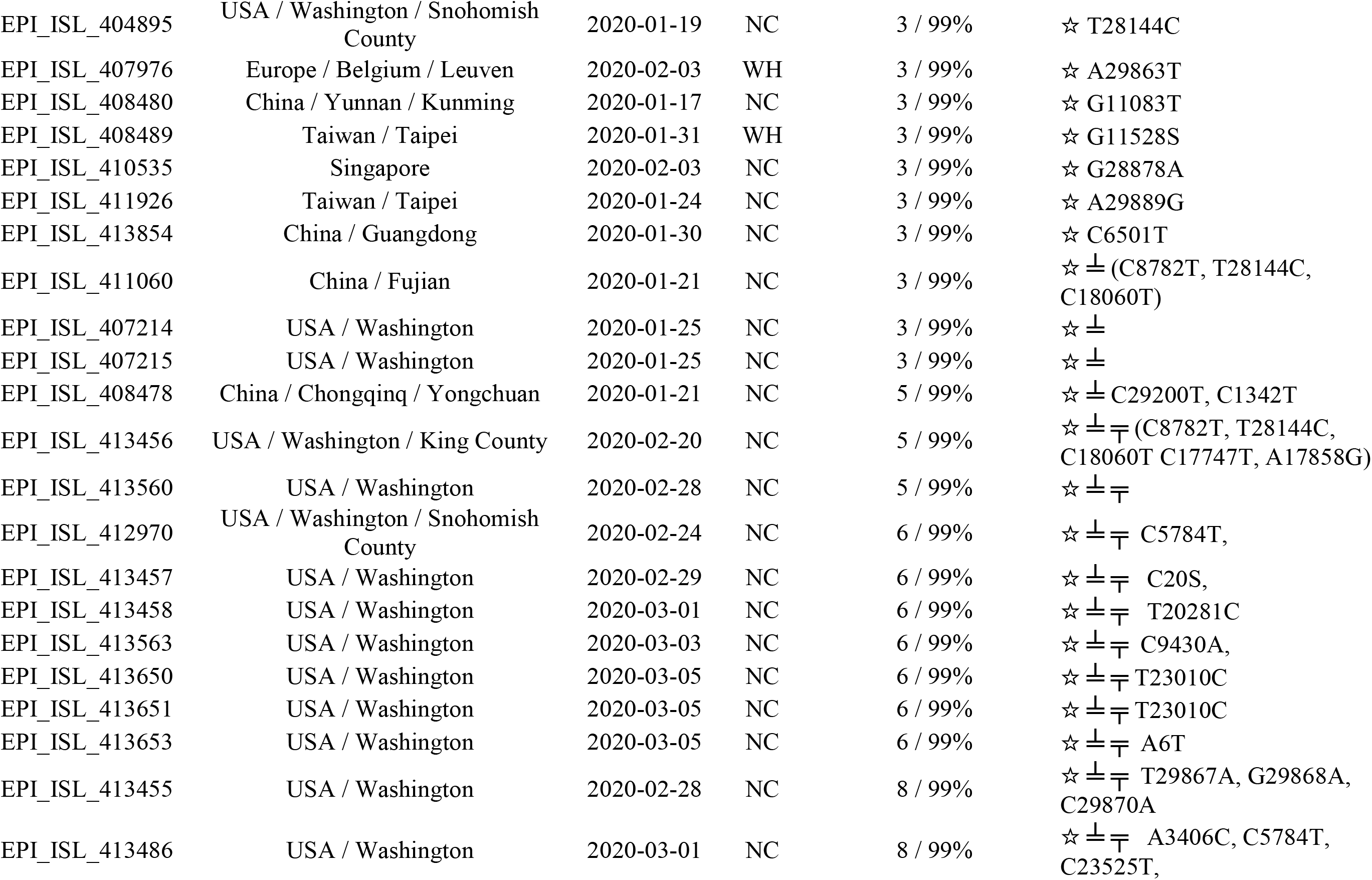
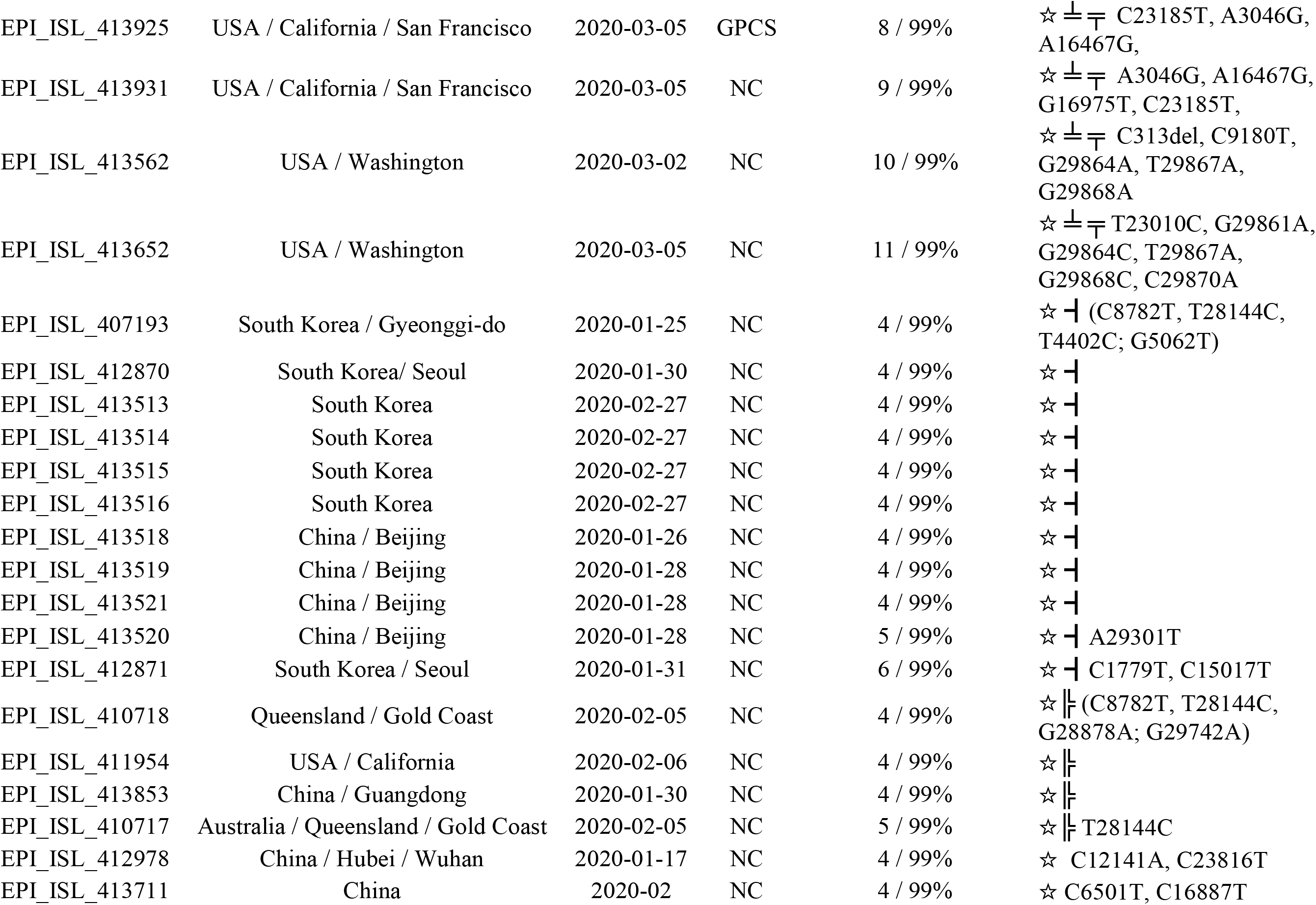
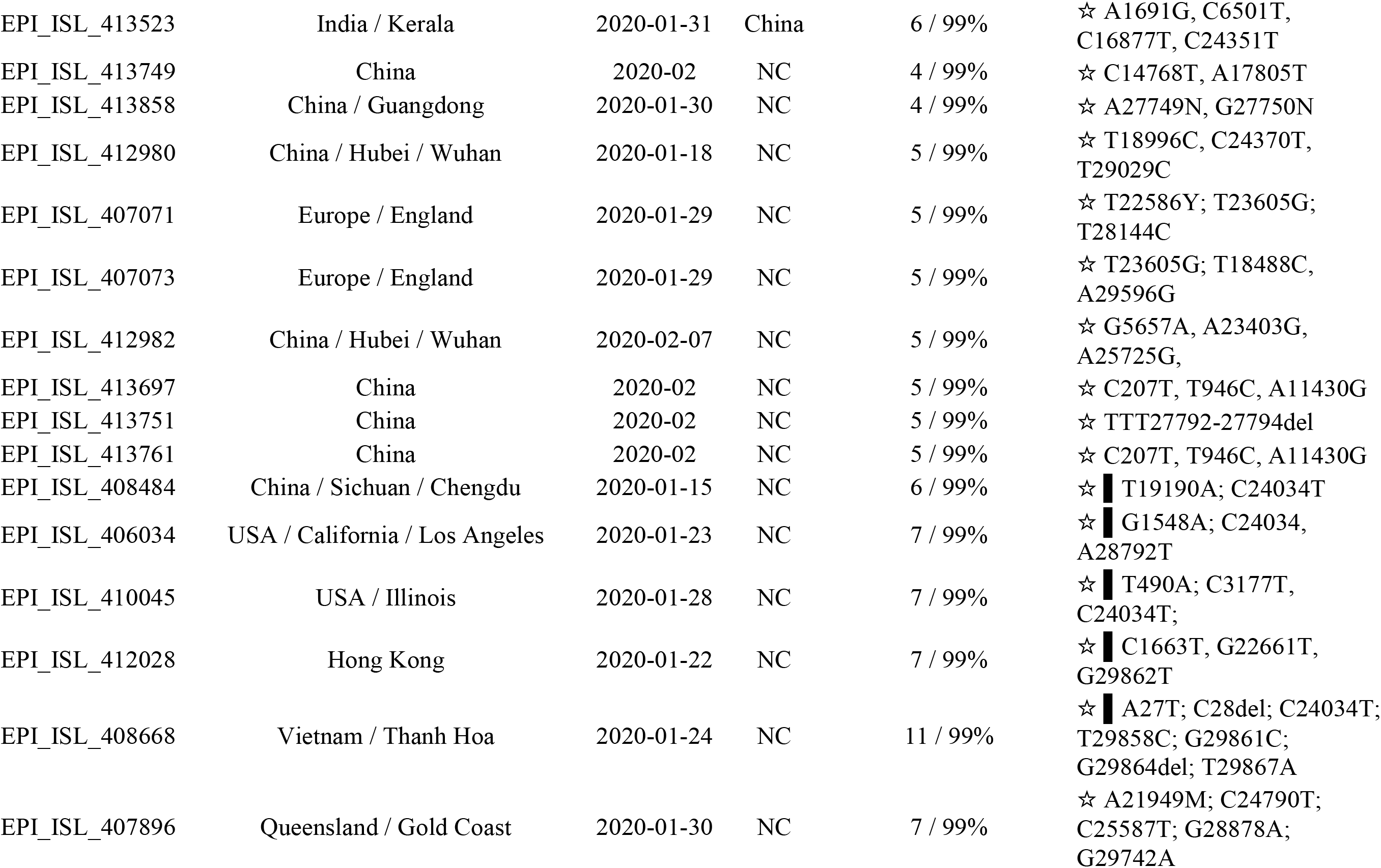
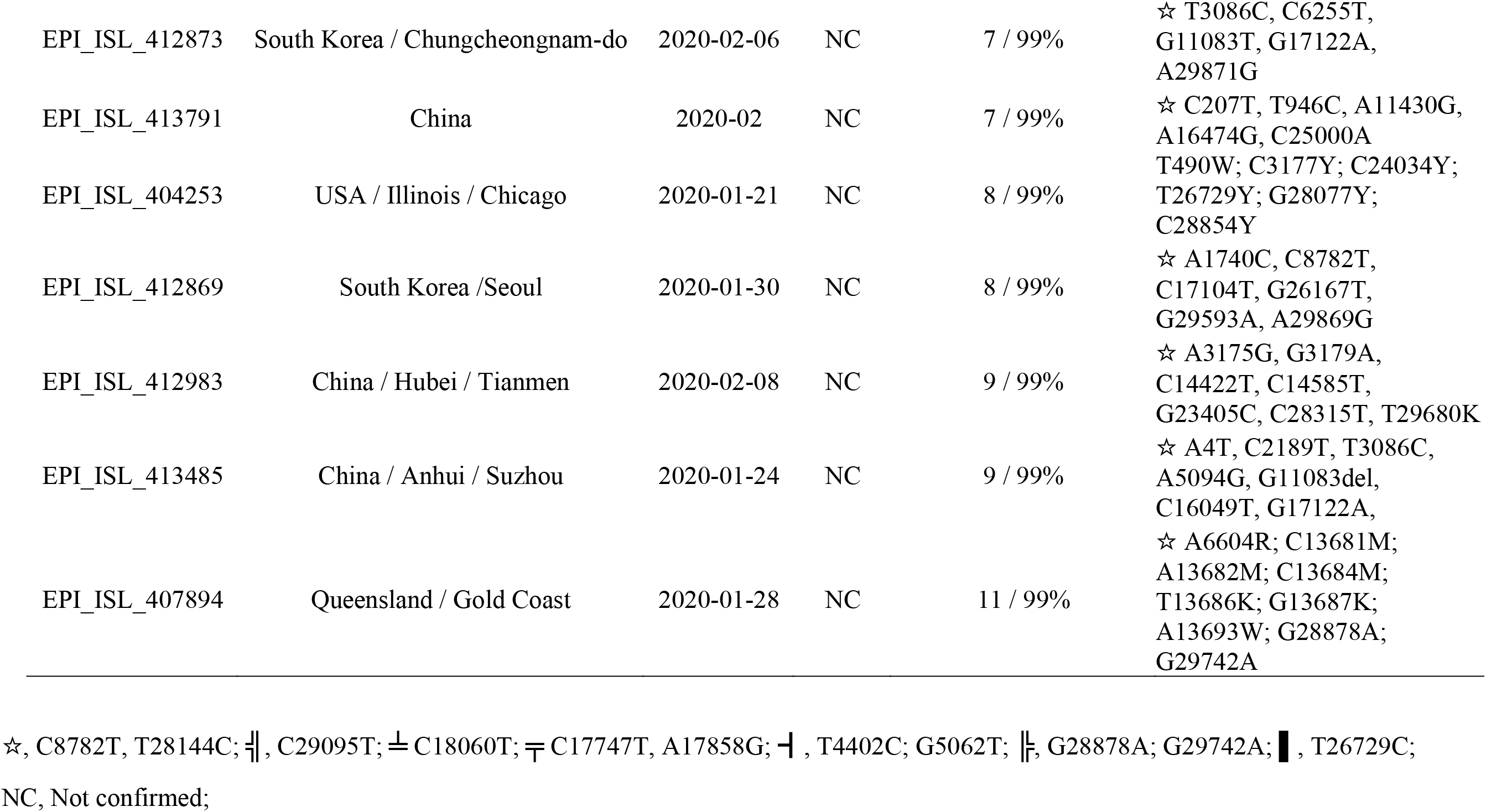
Mutational analysis of genome sequences in super-transmitter cluster 1.

Detailed analysis of mutation profiles of the genome sequences in ST1 enables us to trace the evolution routes of these viruses in specific region. In Washington State, USA, a genome sequence with three mutations, C^18060^T, C^8782^T and T^28144^C, was reported on Jan 25, 2020. A virus carrying these three mutations was reported in Fujian, China on Jan 21, 2020, suggesting that the virus might have originated from Fujian, China (**Table 2**). This virus was then further transmitted in the Washington area and continued to acquire mutations. Twelve genome sequences reported between March 01 - 05, 2020 in the states of Washington and California contained 6 to 11 mutations. The data provided direct evidence of active evolution that results in a large number of mutational changes during the process of transmission of a single virus within a short period of time. In addition, a genome sequence which has two additional mutations when compared to the original virus in this cluster, but were different from those in genome sequences in Washington, was reported in Sichuan, China, suggesting that the same parental virus was also transmitted across China during this period (**Table 2**).

### Evolution and transmission of super-transmitter 2 (ST2)

ST2 carried the signature mutation G^26144^T, which resulted in the G^251^V amino acid substitution in Orf 3 protein of SARS-CoV-2. The first viral genome in this cluster was reported on Jan. 25, 2020 in Australia and a total of 28 out of the 247 (11.2%) sequences tested were found to belong to ST2. The parental virus had acquired different mutations and had been disseminated to various Asian countries, North America (USA), Europe, South America (Brazil) and transmission in Australia. Viruses in this cluster seemed to be extensively transmitted by the end of January and lasted till early February. By the end of February, however, transmission efficiency of such viruses seemed to have dropped, as only 4 oof 28 sequences reported during the period Feb. 26 to Mar. 03. 2020 belong to this cluster. Viruses in this cluster were also found to have significantly mutated. Examples are three genomes reported from Switzerland on Feb. 29, 2020, in which 9, 10 and 11 mutations were identified respectively (**Table 3**). Our data showed that as many as eight additional mutations were acquired by the parental virus within 30 days (from Jan 28 to Feb. 29, 2020), representing a mutation rate of 3.3e-3 subs/site/year (29kb genome size), which was much higher than the predicted mutation rate of SARS-CoV-2 (8.0e-4 subs/site/year).

**Table 3.**
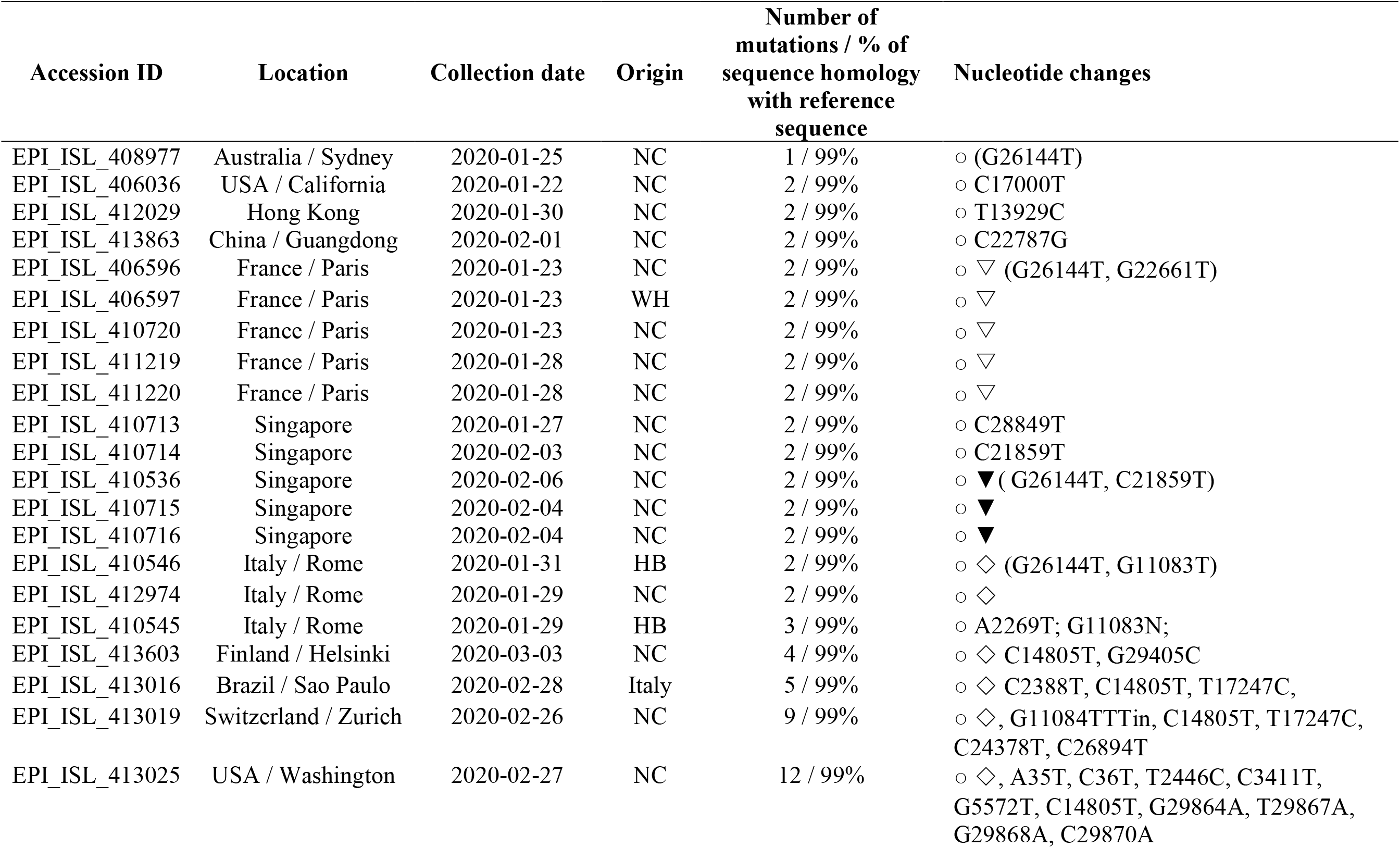
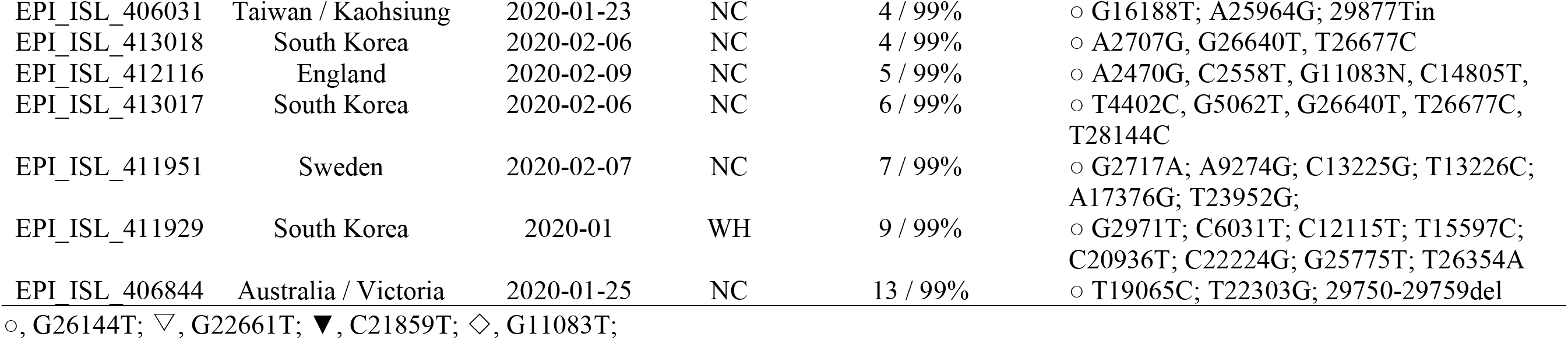
Mutational analysis of genome sequences in super-transmitter cluster 2.

### Evolution and transmission of super-transmitter 3 (ST3)

ST3 carried the signature mutation G^11083^T, which caused the L^3606^F amino acid substitution in the Orf 1 protein of SARS-CoV-2. The first viral genome in this cluster was reported on Jan. 18, 2020 in Chongqing, China and a total of 22 (9%) genome sequences were reported so far. It has since been transmitted to some Asian countries including Singapore, as well as Japan, Europe, USA and Australia (**Table 4**). Like ST1 and ST2, viruses in this cluster were also found to mutate efficiently, with one genome reported on Feb. 27, 2020 from Washington, USA, carrying 12 mutations. Our data showed that a total of eleven mutations were acquired by the parental virus within a 40 days period (from Jan 18 to Feb. 27, 2020), with a mutation rate of 2.8e-3 subs/site/year (29kb genome size), which was again much higher than the predicted mutation rate of SARS-CoV-2 (8.0e-4 subs/site/year). Curiously, there is no virus of this cluster being reported in Iran, a country with one of the highest incidence of SARS-CoV-2 infections. However, two genome sequences from Australia, which belong to viruses recovered from patients with travel history to Iran, were reported, suggesting that this cluster of virus might also contribute to the outbreaks in Iran. In addition, the first genome sequence from Brazil, which might have originated from Italy, also belonged to this cluster (**Table 4**).

**Table 4.**
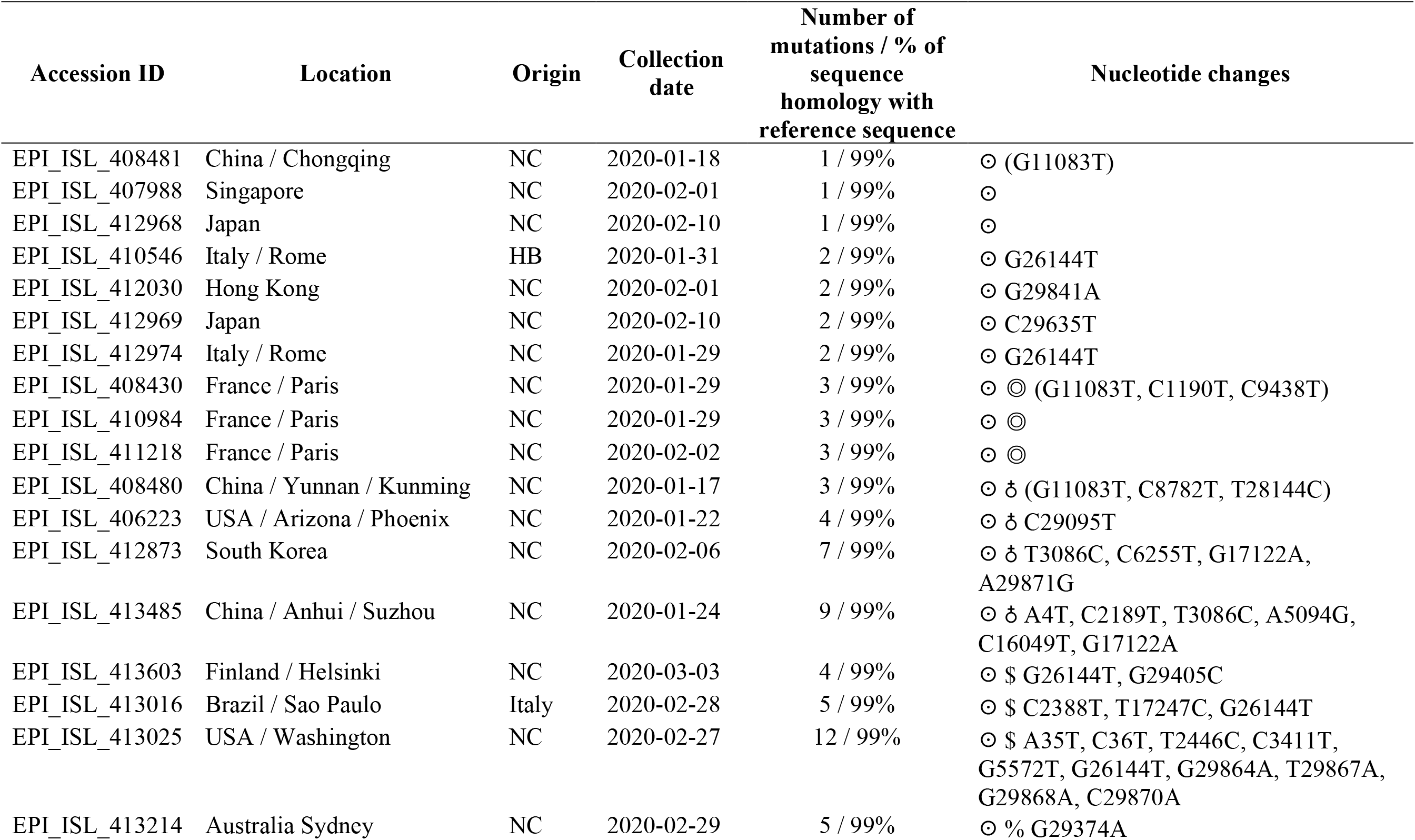
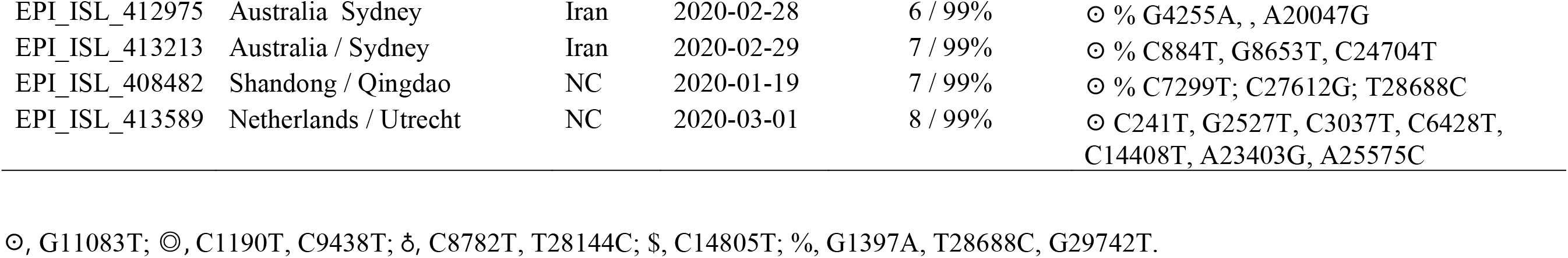
Mutational analysis of genome sequences in super-transmitter cluster 3.

### Evolution and transmission of super-transmitter 4 (ST4)

ST4 carried a signature mutation profile that consists of three mutations: C^241^T, C^3037^T and A^23403^G. The C^241^T and C^3037^T changes are silent mutations, whereas A^23403^G results in the D^614^G substitution in the spike (S) protein of SARS-CoV-2. ST4 viruses were found to be transmitted only in Europe, with the exception of one genome from Mexico with travel history from Italy, and contributed to the current explosive increase in incidence of COVID-19 in Europe (**Table 5**). Genomes in ST4 were reported more recently, from the end of February to early March. A total of 21 out of 247 (8.4%) genome sequences have been reported so far. Among the 247 genome sequences of the four clusters of super-transmitters, there is no genome containing either one of these three mutations or a combination of two of these three mutations, suggesting that the parental viral genome of ST4 could not be identified. The first virus of this cluster was reported on Jan 28, 2020 in Germany. This virus acquired another silent mutation, C^14408^T, and further spread to other countries in Europe. This virus was also found to mutate efficiently, with three genomes reported on Feb. 29, 2020 from Switzerland carrying 9, 10 and 11 mutations respectively. Eight additional mutations were acquired by the parental virus within 30 days (from Jan 28 to Feb. 29, 2020), with a mutation rate of 3.3e-3 subs/site/year (29kb genome size), which was much higher than the predicted mutation rate of SARS-CoV-2 (8.0e-4 subs/site/year). At a later stage, ST4 viruses became the most efficient in causing transmission in Europe. Among the 28 genomes reported between Feb. 20 to March 03, 2020, 20 (71%) belonged to this transmitter (**Table 5**).

**Table 5.**
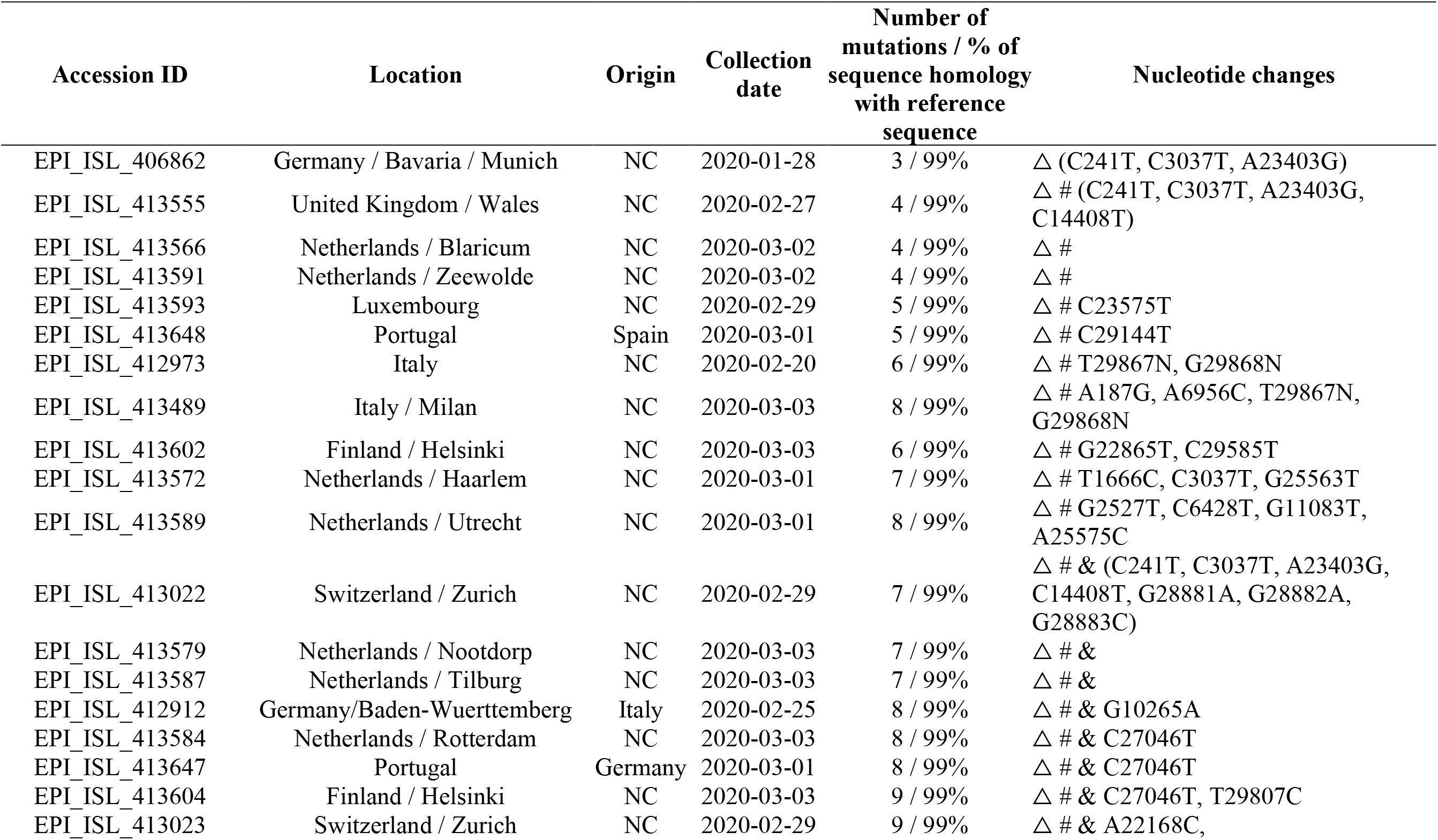
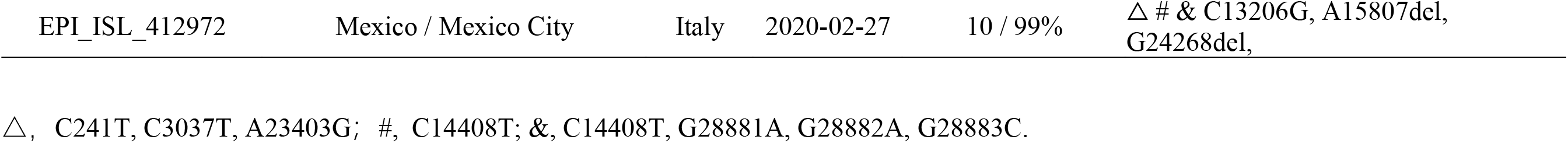
Mutational analysis of genome sequences in super-transmitter cluster 4.

### Temporal and spatial distribution of super-transmitters of SARS-CoV-2

To better understand the temporal and spatial distribution of these super-transmitters, we plot variation in the types of genome sequences recovered from different continents against time. The original viruses were transmitted in the first week before the emergence of these super-transmitters. ST1 was the first batch of viruses that emerged and dissemination continued throughout the study period. Other STs emerged at different time points and transmission also peaked at different dates. Transmission of ST2 and ST3 mainly occurred between mid January to mid February. Transmission of ST4 viruses mainly began at the end of February. Viruses of the four clusters exhibited much higher mutation rate than those which exhibited diverse genetic profiles and could not be allocated into specific genetic cluster when compared to the original genome (**Fig 3a**). ST1 viruses were those which were disseminated extensively in China, in particular in the later stage of the outbreak (**Fig 3b**). ST1, ST2 and ST3 were prevalent in other Asian countries (**Fig 3c**). All the four clusters were involved in the outbreaks in Europe in the early stage, but ST4 was the cluster that eventually transformed the outbreaks in Europe to the pandemic level (**Fig 3d**). In Oceania, ST1 was involved mainly in the early stage of outbreak, yet ST2 became dominant at the later stage (**Fig 3e**). ST1 and other types of viruses were the major transmitters in the US. ST1 was shown to be transmitted mainly in the states of Washington and California, whereas the other types were mainly transmitted in other states, (**Fig 3f, Table S1**).

**Figure 3.**
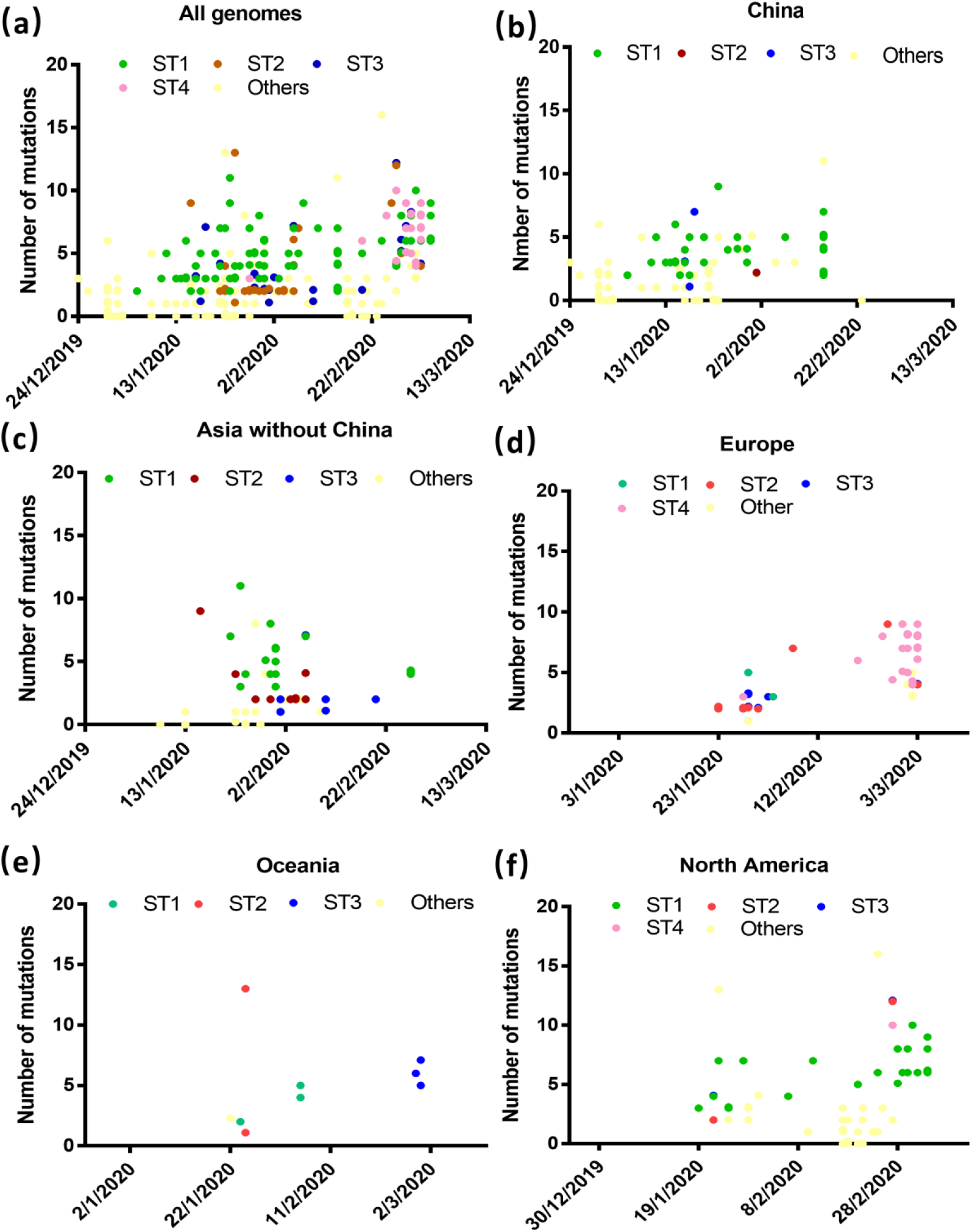
Changes in the distribution pattern and mutation rate of different super-transmitter clusters in various continents over time. Distribution of different STs and their mutations (a) Overall, (b) in China, (c), in Asian countries excluding China, (d) in Europe, (e) in Oceania, and (f) in North America. Two genomes with over 20 mutations were not included to facilitate easy visualization of the graphs.

**Figure 4.**
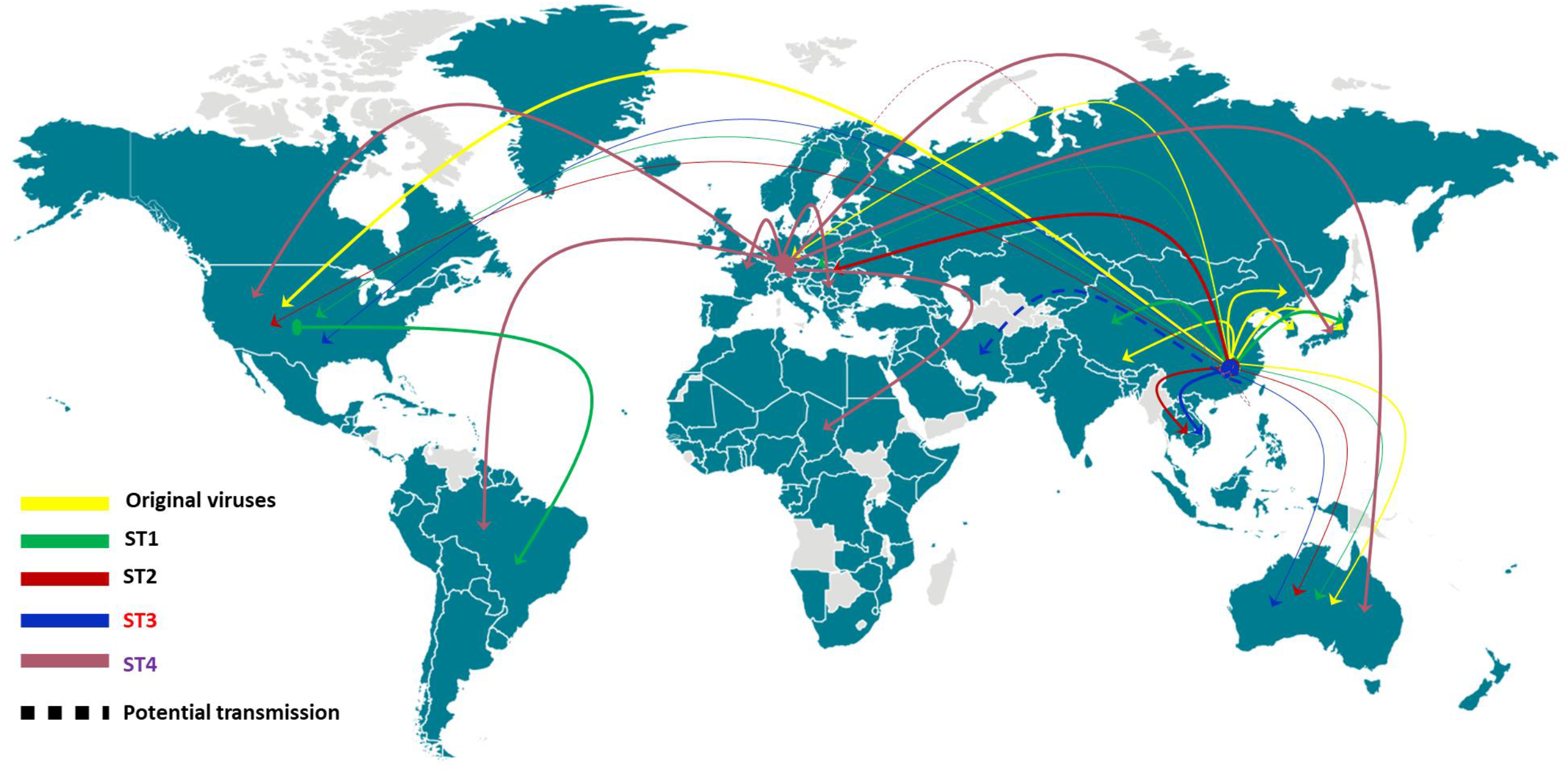
Transmission of super-transmitters and other derivatives of the original SARS-CoV-2 in different areas of world. The derivatives of the original virus have been transmitted worldwide and contributed to the early outbreak of COVID-19. ST1 transmits mainly in Asia and the US but was less prevalent in other parts of the world. ST2 and ST3 was transmitted mainly in Asian countries other than China, as well as Europe from mid of January to mid of February. ST4 was transmits mainly in Europe in the beginning and then transmitted to all over the world.

### Distribution of different types of most recent SARS-CoV-2 in different parts of the world

Upon finishing our manuscript, we went to check the available genome sequences in the database and found a rapid increase of numbers of sequences. A total of 1539 genome sequences reported after February 29, 2020 were included for a quick analysis to identify the type of these most recent genomes. As shown in Table 6, most of the genomes were reported from USA (968 / 63%) and Europe (441 / 29%), where the pandemics were the most server. It is good to see some genomes from Africa (20 / 1%) and South America (23 / 2%), which were minimally reported before March 01, 2020. Among these genomes, 89% of the genomes belonged to ST1-4 with ST4 being the most dominant (56%), while the original derivatives accounted for only 11%, which were mainly reported in UK and Netherland. In Africa, ST4 (18/20, 90%) was the major type with some of the cases showing travel history to Europe; in Asia, the major types became ST3 (17/33, 52%) and ST4 (16/33, 48%); in Europe, all types were presence with ST4 being the dominant one (668/968, 69%); all the types were reported in the US with ST1 (282/441, 62%) and ST4 (137/441, 14%) being the dominant; in Canada, all types except for ST1 were present; in Oceania, all four STs were present with ST3 being the dominant; in South America, ST1 and ST4 were the most dominant types (**Table 6**).

**Table 6.** Distribution of different types of SARS-CoV-2 over the world after February 29, 2020.

| Types of viruses | Numbers of genomes (%) | Major areas | Locations / numbers of genomes (%) |  |  |  |  |  |  |
| --- | --- | --- | --- | --- | --- | --- | --- | --- | --- |
|  |  |  | Africa | Asia | Europe | NA (USA) | NA (Canada) | Oceania | South America |
| **Total** | 1539 |  | 20(1) | 33(2) | 968(63) | 441(29) | 11(1) | 27(2) | 23(2) |
| **Original derivatives** | 173 (11) | Europe (UK and Netherland) | 0(0) | 0(0) | 158(16) | 11(3) | 2(2) | 0(0) | 2(9) |
| **ST1** | 340 (21) | USA, Europe, South America | 1(5) | 1(3) | 30(3) | 282(64) | 0(0) | 5(19) | 8(35) |
| **ST2** | 172 (11) | Europe | 1(5) | 0(0) | 111(12) | 9(2) | 2(2) | 3(11) | 0(0) |
| **ST3** | 132 (9) | Asia, Europe | 0(0) | 17(52) | 119(12) | 12(3) | 5(5) | 15(56) | 0(0) |
| **ST4** | 856 (56) | Africa, Asia, Europe, Oceania | 18(90) | 16(48) | 663(69) | 137(14) | 4(4) | 5(19) | 12(52) |

## Discussion

We conducted detailed and comprehensive analyses of sequences of SARS-CoV-2 reported from December to March 05, 2020 and deposited in the GISAID database. The detailed analysis of 247 high quality genome sequences of SARS-CoV-2 provides insight into the evolution and transmission of this novel virus (**Fig 5**). The ancestor of SARS-CoV-2 could have emerged at a date as early as November, 2019 based on results of phylodynamics analysis of these genome sequences. According to the time line of outbreaks, the original virus from Wuhan city and HNSM was responsible for the widespread transmission of SARS-CoV-2 in different parts of the world in January. The origin of the outbreak was not limited to HNSM, those which occurred in multiple sites in Wuhan city might have contributed to the early transmission events and subsequent dissemination to different parts of China and various countries around the world. These data also implied that wild animals in HNSM may not be the intermediate host of SARS-CoV-2 as sources other than HNSM are also considerd the origin of this virus. Given the fact that multiple patients in Wuhan were simultaneously infected by viruses of different genetic composition in the initial outbreak, we hypothesize that a common wild animal such as wild rat would be the most likely intermediate host. Alternatively, a common environmental factor, such as a faulty sewage system, may be involved. It is therefore necessary to investigate the role of a common animal vector or dissemination route in eliciting the SARS-CoV-2 outbreak.

Interestingly, as the original virus continued to transmit in China and over the world, it has evolved into four major genetic clusters, namely super-transmitter clusters, along with other non-cluster variants derived from the original virus. Each ST cluster carried its unique signature mutation(s), which enable us to trace its origin and transmission dynamics. In the early transmission stage (December and January, 2020), variants from the original virus were dominant, yet by the end of February and early March, these four super-transmitters became dominant, with different STs being prevalent in different regions of the world. ST1 was prevalent in China and other parts of Asia and became the major virus that caused severe outbreaks in Washington and California states in the US and South Korea; ST2 and ST3 were extensively transmitted in other parts of Asia and Europe during the end of January and early February but its prevalence dropped at the end of February and early March, and was replaced by ST4 which contributed to the pandemic in Europe. Mapping the mutational profile of viral genomes enables us to trace the transmission of viruses of different clusters in different parts of the world. Interestingly, ST4 was not reported in China or other parts of the world. The first genome of this cluster was reported in Germany and contributed to the rapid dissemination of SARS-CoV-2 in Europe. Mutation profile with ST4 is unique, with three mutations being observed in the first viral genome. Genomes with one or two of these three mutations were not reported anywhere. These data do not simply imply that ST4 originated from Europe. One limitation of the study is that we can only utilize currently available genome sequences. The lack of genome sequence of ST4 in other continent does not necessarily mean that ST4 viruses are not present in other continents. A second limitation of this study is the lack of data to explain the mechanisms underlying the evolution of these genetic clusters into super-transmitters. Every ST cluster carried at least one amino acid mutation in different protein. Whether such mutational changes were the key step that enabled SARS-CoV-2 to evolve into super-transmitters must be investigated in future research studies.

Lastly, our data also provided insight into the major transmitting viruses in current pandemic areas in the world. For example, in Italy, ST2, ST3 were reported in the end of January, while ST4 was reported in February and early March. Similar trends were seen in other countries with exception that a higher proportion of the original viral genomes were reported in Netherland. In the US, the original viruses were reported in other states, while ST1 was the major virus that caused outbreak in Washington and California States. Other ST genomes were also sporadically reported in the US. Although data from Iran is not available, two genomes reported from Australia with travel history from Iran were shown to belong to ST3, suggesting that this cluster was responsible for the pandemic in Iran. In Australia, all genomes were reported except ST4.

Using the signature mutations as markers for different STs, we were able to analysed 1539 genomes reported in March. The data further confirmed that four STs became dominant in March with around 90% of the genomes belonging to these four STs, among which ST4 became the dominant cluster transmitting in Europe. It also stared transmitting to other parts of world including Africa, Asia, North America and Oceania. ST1 was still the major type transmitting in the US and has transmitted to South America in particular Brazil. These data confirmed that ST1 and ST4 would become worldwide transmitter and dominate the future transmission of SARS-CoV-2 in the world.

In conclusion, this study show that four major genetic clusters of viruses evolved from the original SARS-CoV-2 and have transmitted extensively over the world, each becoming dominant in different parts of the world, and that viruses without any signature mutation of the four super-transmitters appear to be transmitted much less efficiently. These super-transmitters exhibit not only high transmission efficiency, but also high mutation rate without compromising infectivity, compromising effectiveness of current infection control effort. Our findings therefore provide important insight into the molecular features of the highly transmissible variants of SARS-CoV-2.

## Data Availability

Genome sequences of SARS-CoV-2were downloaded from the GISAID platform.

## Acknowledgments

We are grateful for the discussion and comments from Prof. Mengsu Yang from City University of Hong Kong. We acknowledge the use of the genome sequences in the GISAID platform. This study was not supported by any research grant.

